# Artificial intelligence-guided detection of under-recognized cardiomyopathies on point-of-care cardiac ultrasound: a multi-center study

**DOI:** 10.1101/2024.03.10.24304044

**Authors:** Evangelos K. Oikonomou, Akhil Vaid, Gregory Holste, Andreas Coppi, Robert L. McNamara, Cristiana Baloescu, Harlan M. Krumholz, Zhangyang Wang, Donald J. Apakama, Girish N. Nadkarni, Rohan Khera

## Abstract

**Background:** Point-of-care ultrasonography (POCUS) enables cardiac imaging at the bedside and in communities but is limited by abbreviated protocols and variation in quality. We developed and tested artificial intelligence (AI) models to automate the detection of underdiagnosed cardiomyopathies from cardiac POCUS.

**Methods:** In a development set of 290,245 transthoracic echocardiographic videos across the Yale-New Haven Health System (YNHHS), we used augmentation approaches and a customized loss function weighted for view quality to derive a POCUS-adapted, multi-label, video-based convolutional neural network (CNN) that discriminates HCM (hypertrophic cardiomyopathy) and ATTR-CM (transthyretin amyloid cardiomyopathy) from controls without known disease. We evaluated the final model across independent, internal and external, retrospective cohorts of individuals who underwent cardiac POCUS across YNHHS and Mount Sinai Health System (MSHS) emergency departments (EDs) (2011-2024) to prioritize key views and validate the diagnostic and prognostic performance of single-view screening protocols.

**Findings:** We identified 33,127 patients (median age 61 [IQR: 45-75] years, n=17,276 [52·2%] female) at YNHHS and 5,624 (57 [IQR: 39-71] years, n=1,953 [34·7%] female) at MSHS with 78,054 and 13,796 eligible cardiac POCUS videos, respectively. An AI-enabled single-view screening approach successfully discriminated HCM (AUROC of 0·90 [YNHHS] & 0·89 [MSHS]) and ATTR-CM (YNHHS: AUROC of 0·92 [YNHHS] & 0·99 [MSHS]). In YNHHS, 40 (58·0%) HCM and 23 (47·9%) ATTR-CM cases had a positive screen at median of 2·1 [IQR: 0·9-4·5] and 1·9 [IQR: 1·0-3·4] years before clinical diagnosis. Moreover, among 24,448 participants without known cardiomyopathy followed over 2·2 [IQR: 1·1-5·8] years, AI-POCUS probabilities in the highest (vs lowest) quintile for HCM and ATTR-CM conferred a 15% (adj.HR 1·15 [95%CI: 1·02-1·29]) and 39% (adj.HR 1·39 [95%CI: 1·22-1·59]) higher age- and sex-adjusted mortality risk, respectively.

**Interpretation:** We developed and validated an AI framework that enables scalable, opportunistic screening of treatable cardiomyopathies wherever POCUS is used.

**Funding:** National Heart, Lung and Blood Institute, Doris Duke Charitable Foundation, BridgeBio

**Research in Context:** 

**Evidence before this study:** Point-of-care ultrasonography (POCUS) can support clinical decision-making at the point-of-care as a direct extension of the physical exam. POCUS has benefited from the increasing availability of portable and smartphone-adapted probes and even artificial intelligence (AI) solutions that can assist novices in acquiring basic views. However, the diagnostic and prognostic inference from POCUS acquisitions is often limited by the short acquisition duration, suboptimal scanning conditions, and limited experience in identifying subtle pathology that goes beyond the acute indication for the study. Recent solutions have shown the potential of AI-augmented phenotyping in identifying traditionally under-diagnosed cardiomyopathies on standard transthoracic echocardiograms performed by expert operators with strict protocols. However, these are not optimized for opportunistic screening using videos derived from typically lower-quality POCUS studies. Given the widespread use of POCUS across communities, ambulatory clinics, emergency departments (ED), and inpatient settings, there is an opportunity to leverage this technology for diagnostic and prognostic inference, especially for traditionally under-recognized cardiomyopathies, such as hypertrophic cardiomyopathy (HCM) or transthyretin amyloid cardiomyopathy (ATTR-CM) which may benefit from timely referral for specialized care.

**Added value of this study:** We present a multi-label, view-agnostic, video-based convolutional neural network adapted for POCUS use, which can reliably discriminate cases of ATTR-CM and HCM versus controls across more than 90,000 unique POCUS videos acquired over a decade across EDs affiliated with two large and diverse health systems. The model benefits from customized training that emphasizes low-quality acquisitions as well as off-axis, non-traditional views, outperforming view-specific algorithms and approaching the performance of standard TTE algorithms using single POCUS videos as the sole input. We further provide evidence that among reported controls, higher probabilities for HCM or ATTR-CM-like phenotypes are associated with worse long-term survival, suggesting possible under-diagnosis with prognostic implications. Finally, among confirmed cases with previously available POCUS imaging, positive AI-POCUS screens were seen at median of 2 years before eventual confirmatory testing, highlighting an untapped potential for timely diagnosis through opportunistic screening.

**Implications of all available evidence:** We define an AI framework with excellent performance in the automated detection of underdiagnosed yet treatable cardiomyopathies. This framework may enable scalable screening, detecting these disorders years before their clinical recognition, thus improving the diagnostic and prognostic inference of POCUS imaging in clinical practice.

## INTRODUCTION

Point-of-care ultrasonography (POCUS) enables cardiac evaluation as a direct extension of the physical exam,^1^ and is increasingly used as an adjunctive diagnostic tool across communities, outpatient clinics, emergency departments (ED), and inpatient facilities.^2^ However, in contrast to standard transthoracic echocardiography (TTE), POCUS studies are acquired by clinicians in busy clinical settings and for specific diagnostic tasks. Consequently, only a limited number of distinct views are obtained, and image quality and anatomical accuracy are often limited due to a range of environmental factors (equipment and time constraints), patient characteristics (inability to reposition, distress, individual disease states, and anatomy), and operator experience.^3^ Therefore, the videos generated are rarely used beyond addressing acute medical questions.

With the expanding use of handheld technology and software that assists even novice operators in acquiring key cardiac views,^4^ there is a growing realization that POCUS imaging represents a missed window for detecting potentially modifiable, subclinical cardiac diseases. Therefore, even if the intention of the study is quantifying left ventricular function or ruling out acute, life-threatening conditions, acquired POCUS data may enable the opportunistic screening of underdiagnosed cardiac disorders. This applies especially to cardiomyopathies that have a long presymptomatic course and are increasingly treatable with disease-modifying therapies, such as hypertrophic cardiomyopathy (HCM)^5^ or transthyretin amyloid cardiomyopathy (ATTR-CM).^6^ This is critical as only a minority (10-20%) of HCM or ATTR-CM cases are identified clinically.^8–11^ Furthermore, the reliance on serial multi-modality imaging exacerbates disparities in diagnosis that disproportionately affect traditionally disadvantaged groups.^12–14^ In recent years, echocardiography has benefited from advances in computer vision and medical artificial intelligence (AI) for automated cardiovascular diagnosis,^15–20^ with improved accuracy relative to standard workflows.^21^ Nevertheless, such AI algorithms are almost invariably developed and validated using videos acquired by expert technicians and interpreted by board-certified readers. Furthermore, across several settings, access to standard echocardiography remains limited and prone to referral and selection bias. POCUS studies are increasingly done across high- and low-resource settings for both cardiovascular and non-cardiovascular complaints. Thus, they represent an untapped yet accessible and scalable resource for opportunistic screening.

Here, we propose and implement a framework for POCUS-adapted video-based AI models, demonstrating their ability to efficiently detect under-diagnosed cardiomyopathies from real-world POCUS studies acquired over a decade across the emergency departments of two large hospital systems. Our approach incorporates a range of natural and synthetic augmentation methods to simulate off-axis acquisitions from variable views, thus enabling downstream inference from limited POCUS acquisitions. We then assess the diagnostic and prognostic potential of this technology, offering insights into missed or delayed diagnosis of key cardiomyopathies.

## METHODS

### Study overview

First, we developed a POCUS-adapted pipeline to train video-based deep learning models for the diagnosis of frequently under-recognized cardiomyopathies (HCM, ATTR-CM) among patients in the Yale-New Haven Health System (YNHHS), an integrated healthcare system of five hospitals and affiliated outpatient clinics across Connecticut and Rhode Island (U.S.). To maximize the accuracy of our labels, we required confirmatory assessment by cardiac magnetic resonance (CMR), or cardiac scintigraphy testing, as described below. Second, we deployed the POCUS-adapted models among retrospective cohorts of consecutive independent patients undergoing real-world POCUS imaging across the EDs of YNHHS, as well as a distinct, external health system (Mt Sinai Health System [MSHS], New York) to assess their ability to discern HCM and ATTR-CM cases and prognosticate long-term outcomes (**Figure 1**).

**Figure 1.**
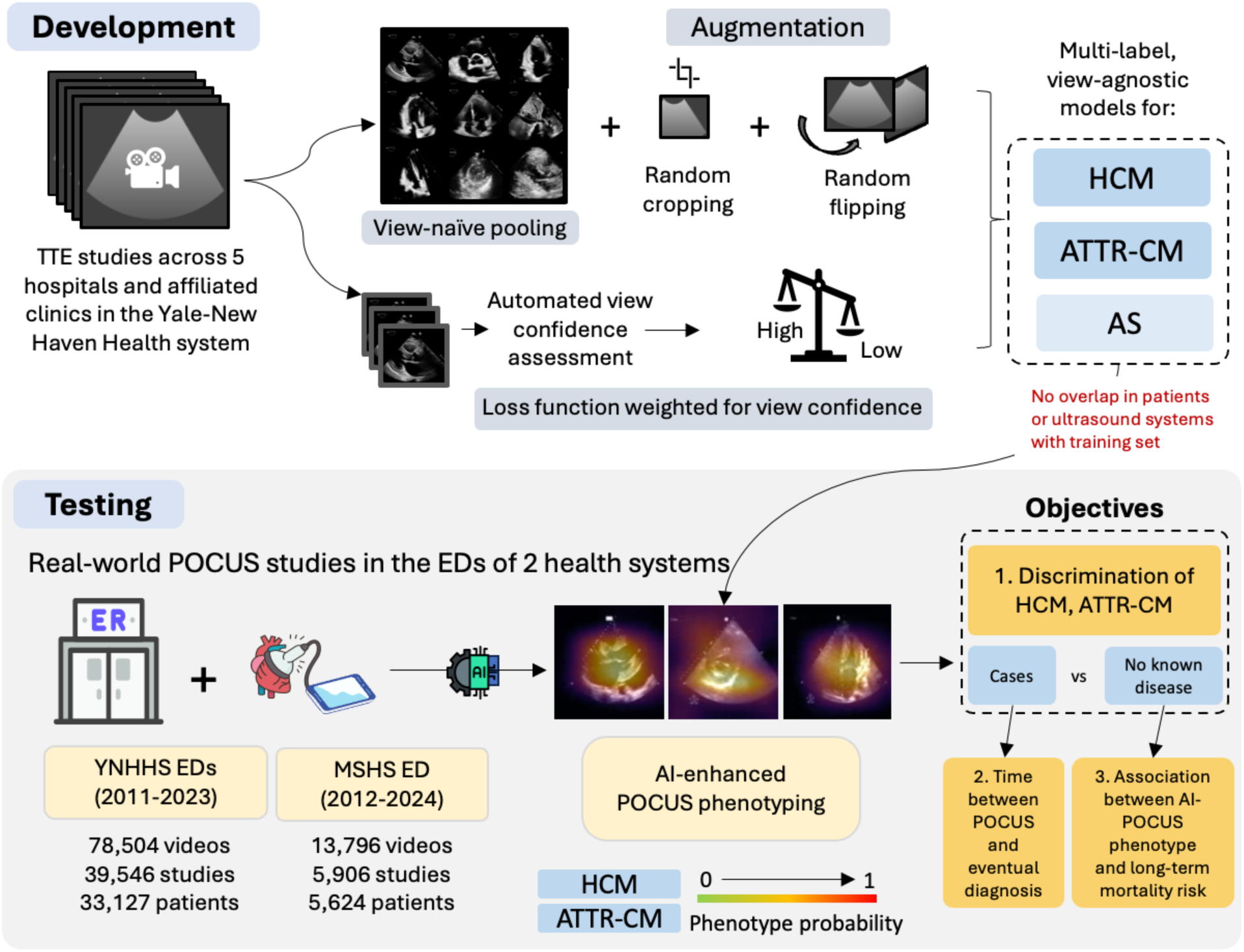
Study overview. Overview of study design and datasets. AI: artificial intelligence; AS: (severe) aortic stenosis; ATTR-CM: amyloid transthyretin cardiomyopathy; ED: emergency department; HCM: hypertrophic cardiomyopathy; MSHS: Mt Sinai Health System; POCUS: point-of-care ultrasound; TTE: (standard) transthoracic echocardiography; YNHHS: Yale-New Haven Health System.

### Study objectives

The primary objective of the analysis was to develop and validate the performance of an AI-enabled POCUS-adapted approach in identifying HCM and ATTR-CM among individuals undergoing real-world POCUS in EDs at two large and diverse health systems.

Acknowledging the expected underdiagnosis of HCM and ATTR-CM, we further addressed the following secondary objectives. First, among patients who had a POCUS and were eventually diagnosed with HCM or ATTR-CM, to evaluate the potential for opportunistic screening in early disease diagnosis, we examined the time difference between their first positive screen by AI-POCUS and their eventual clinical diagnosis. Second, to examine the potential prognostic implications of a positive screen among patients without known HCM or ATTR-CM, we assessed the association between the output probability for each unique label at the time of POCUS and all-cause mortality.

### Methodological Approach

#### Development of a multi-label disease classifier (YNHHS; TTE development cohort) Data access

We queried the YNHHS electronic health record (EHR) and the linked echocardiographic repository for TTEs performed until 12/31/2022 in patients with HCM, ATTR-CM, and controls. We chose these conditions due to their indolent course and delayed diagnosis, overlapping phenotypic features yet distinct underlying pathology, and given that they are not routinely discernible on cardiac POCUS or TTE and often require multi-modality assessment that results in limited access and potential diagnostic delays.

#### Label definitions

We used a composite of diagnostic elements to define distinct disease labels. We first identified all individuals in the YNHHS with at least one ICD (International Classification of Disease) code suggestive of cardiomyopathy and/or clinical heart failure (**Table S1**). We then narrowed down to those who had confirmatory cardiac imaging, including CMR evidence consistent with HCM,^22^ or abnormal bone scintigraphy (with Tc^99m^-pyrophosphate [PYP]) for ATTR-CM (see **appendix pp 2**).^23^ Given that HCM is a genetic cardiomyopathy, we included all available echocardiograms regardless of their timing relative to the time of diagnosis.^8^ For ATTR-CM cases, we defined the time of diagnosis as the time of the positive PYP scan, and, to account for the expected delay between disease onset and diagnosis (reported median delay of ∼12-13 months),^23,24^ we included echocardiograms performed up to 12 months before this date (or any time after). Furthermore, we specifically identified and enriched our sample for cases of severe AS based on the interpretation of a TTE exam by a board-certified reader and in agreement with existing guidelines.^25^ This was done to ensure that the model learned to identify it as a separate pathology, as it is a potential confounder for cardiomyopathies and also associated with LVH. Controls were selected by randomly sampling TTEs from the same period after excluding any positive HCM, ATTR-CM, or AS cases, as well as cases with equivocal CMR or PYP results for HCM or ATTR-CM.

#### Development cohort

We developed our models across all views of TTE from participants who did not contribute any data to the ED POCUS subset, and randomly split our development cohort at the patient level into a derivation (training 75%, validation 15%), and TTE testing (10%) set. This prevented data leakage and ensured that distinct echocardiograms belonging to the same patient would not be present across different sets. To ensure controls did not include missed cases during training, we excluded any echocardiographic study with a measured interventricular septal thickness during diastole (IVSd) of 1·3 cm or greater from the controls of our training set. Critically, this was *not* done during validation or testing to ensure reliable model performance assessment. In total, we identified 10,702 TTEs with 290,245 videos from 8,460 unique patients with HCM, ATTR-CM, and/or AS, as well as controls, to train and validate view quality-adjusted and view-agnostic multi-label video classifiers for each key condition (**Table 1**).

**Table 1.** Summary of cohort characteristics.

|  | <u>Development (TTE) cohort</u> |  | <u>Testing ED cohorts (POCUS)</u> |  |
| --- | --- | --- | --- | --- |
|  | <u>Train/Validation</u> | <u>Testing</u> | <u>YNHHS</u> | <u>MSHS*</u> |
| **Number of unique patients** | 7,621 | 839 | 33,127 | 5,624 |
| **Number of unique studies** | 9,667 | 1,035 | 39,546 | 5,906 |
| **Number of unique videos, including:** | 261,756 | 28,489 | 78,054 | 13,796 |
| *Parasternal long-axis view (PLAX)* | 30,556 (11·7) | 3,283 (11·5) | 15,751 (20·2) | 5,876 (42·6) |
| *PSAX (papillary muscle level)* | 31,779 (12·2) | 3,313 (11·6) | 52,477 (67·2) | 5,237 (38·0) |
| *Apical 4-chamber (A4C)* | 25,528 (9·8) | 2,649 (9·3) | 9,826 (12·6) | 2,683 (19·4) |
| **Participant-level demographics** |  |  |  |  |
| **Age at the time of scan (years)** | 70 [59-79] | 71 [61-80] | 61 [45-75] | 57 [39-71] |
| **Sex (n,%)** |  |  |  |  |
| *Female* | 3,694 (48·5) | 413 (49·2) | 17,276 (52·2) | 1,953 (34·7) |
| *Male* | 3,927 (51·5) | 426 (50·8) | 14,923 (45·0) | 2,470 (44·0) |
| *Unknown* | - | - | 928 (2·8) | 1,201 (21·3) |
| **Hispanic Ethnicity (n, %)** |  |  |  |  |
| *Hispanic* | 489 (6·4) | 50 (6·0) | 5,073 (15·3) | 1,094 (19·5) |
| *Non-Hispanic* | 6,806 (89·3) | 762 (90·8) | 26,796 (80·9) | 2,656 (47·2) |
| *Unknown* | 326 (4·3) | 27 (3·2) | 1,258 (3·8) | 1,874 (33·3) |
| **Race (n, %)** |  |  |  |  |
| *Asian* | 102 (1·3) | 14 (1·7) | 616 (1·9) | 189 (3·4) |
| *Black or African American* | 527 (6·9) | 60 (7·2) | 8,040 (24·3) | 1,385 (24·6) |
| *White* | 6,353 (83·4) | 701 (83·6) | 19,535 (59·0) | 1,215 (21·6) |
| *Other/Unknown* | 639 (8·4) | 64 (7·6) | 4,936 (14·9) | 2,835 (50·4) |
| **Video-level labels** |  |  |  |  |
| *HCM* | 41,602 (15·9) | 4,796 (16·8) | 279 (0·4) | 74 (0·5) |
| *ATTR-CM* | 7,920 (3·0) | 922 (3·2) | 172 (0·2) | 32 (0·2) |
| *AS\*\** | 37,270 (14·2) | 4,195 (14·7) | 1,130 (1·4) | 584 (4·1) |

Values are presented as median [25<sup>th</sup>-75<sup>th</sup> percentile] or counts (percentages). \*Diagnoses in the MSHS cohort were adjudicated based on ICD (International Classification of Diseases) codes. \*\*Aortic stenosis (AS) refers to severe AS by echocardiography in the YNHHS dataset, or any AS by diagnosis codes in MSHS. *AS*: aortic stenosis; *ATTR-CM*: transthyretin amyloid cardiomyopathy; *ED*: emergency department; *HCM*: hypertrophic cardiomyopathy; *MSHS*: Mt Sinai Health System; *POCUS*: point-of-care ultrasound; *YNHHS*: Yale-New Haven Health System.

### Testing (POCUS) cohorts (YNHHS, MSHS)

We reviewed all available cardiac POCUS studies performed by an ED provider across the independent internal testing cohort from YNHHS (2013-2023) and an independent external cohort from MSHS (2012-2024). Studies were generally performed using compact mid-range ultrasound systems created for point-of-care, ED and operating room use (i.e., Sparq Ultrasound system, Philips, in YNHHS) and were not interpreted by cardiologists. We excluded any POCUS studies performed among those with end-stage renal disease, transplanted heart, and/or prosthetic aortic valve (**Table S1**), as well as POCUS studies that exclusively included non-cardiac imaging (i.e., lung ultrasound) with none of the major cardiac views typically captured in a cardiac POCUS (at least one of parasternal long axis [PLAX], parasternal short axis at the papillary muscle level [PSAX] and/or apical four-chamber [A4C] views). In the YNHHS cohort, our approach resulted in 39,546 studies with 78,054 eligible POCUS videos from 33,127 unique patients. In the MSHS cohort, we identified 5,906 studies with 13,796 eligible POCUS videos from 5,624 unique patients. Labels in YNHHS were adjudicated as in the original development cohort (for AS, we required a TTE showing severe AS up to 12 months after the scan). Patient outcomes were extracted through the EHR, which is linked to in-hospital and out-of-hospital death data. In MSHS, the presence of HCM, ATTR-CM, and AS was defined using specific ICD codes (see **Table S1**).

### Automated view characterization and alignment assessment

We implemented our previously described end-to-end pre-processing pipeline that loads, deidentifies, and pre-processes echocardiographic videos (further described in the **appendix pp 2-3**).^19^ On these preprocessed videos, representing different views acquired during a TTE, we applied a validated CNN that enables video-level classification of 18 standard echocardiographic views by assigning a probability that a given video corresponds to a standard anatomical view (with probabilities adding up to 1 across all views).^26^ This was done both for TTE studies, as well as for POCUS studies in order to identify acquisitions that matched standard echocardiographic views given the lack of standardization seen in point-of-care imaging. The highest probability value ((0-1]) for each view was used to define the most likely view, and we used the value of the probability as a surrogate metric of confidence in the anatomical alignment relative to standard echocardiographic views, with probabilities ≥0.5 considered suggestive of high confidence.

### Designing a view-agnostic training pipeline adapted for low-quality acquisitions

We designed a training framework that integrated multiple views (apical, parasternal long, parasternal short, and subcostal views) without annotations and a customized training loss to assign higher weights to low-quality, off-axis videos from participants positive for the labels of interest. We first initialized a 3D-ResNet18 CNN architecture by using pre-trained weights from the Kinetics-400 dataset,^27^ and further modified the output layer of the label to enable multi-label classification for HCM, ATTR-CM, and severe AS. To reflect the unique challenges of screening for relatively rare cardiomyopathies on POCUS scans, we implemented a range of customizations, as follows:

#### Natural and synthetic data augmentation methods

We trained (a) separate models for each key view-of-interest, namely PLAX, PSAX, and A4c, as well as (b) all-inclusive, view-naïve models trained in pooled datasets that included all parasternal (long and short) and apical views with the classifier blinded to the input view. This enabled a head-to-head comparison of how view-specific versus view-agnostic approaches generalize to real-world POCUS acquisitions. We further applied a series of data augmentations to account for variable orientation and off-axis views, including random horizontal flipping and rotation (**appendix pp 3-4**).

#### Quality-adjusted weights and loss function

We customized our loss function to favor inference from acquisitions where the view-classifier had lower confidence in assigning a standard view. For this, the sigmoid binary cross entropy loss function was adapted to incorporate label-specific weights to account for rare labels and inverse weighting based on the view alignment probabilities (**appendix pp 4-5**).^26^ The aim was to penalize the model for missing under-represented labels, especially in a challenging view.

#### Model training and inference

All models were trained for a maximum of 30 epochs with early stopping, such that if the mean validation area under the receiver operating characteristic (AUROC) on the validation cohort for the three labels of interest (HCM, ATTR-CM, AS) did not improve for 5 consecutive epochs, training was terminated and the weights from the epoch with maximum validation AUROC were used for final evaluation. Details of training hyperparameters and the computational framework are provided in the appendix (**appendix pp 3-4**). At the time of inference, we averaged four 16-frame-clip-level predictions to obtain video-level predictions for each label. We analyzed the performance of video- and study-level predictions by averaging class-specific probabilities from all videos acquired during the same study.

### Model explainability

To assist with explainability, we generated sample saliency maps for the most confident cases using Gradient-weighted Class Activation Mapping (Grad-CAM).^28^ We present the pixel-wise maximum along the temporal axis to capture the most salient spatial regions as a heatmap overlayed on the videos leveraged in defining the respective conditions.

### Statistical analysis

Continuous variables are presented as median [25^th^-75^th^ percentile] and compared using the Mann-Whitney across two groups. Categorical variables are summarized as counts (and percentages) and compared across distinct groups using the χ^2^ test. Metrics of discrimination (i.e., area under the receiver operating characteristic curve [AUROC]) for each label and the estimated differences in these metrics between competing models (δ[AUROC]) were computed with corresponding 95% confidence intervals (CI) derived from bootstrapping with 200 replications. Label-specific thresholds were computed individually for HCM, ATTR-CM, and AS, based on the cutoff values for 90% sensitivity in the held-out test set of the development (TTE) cohort. For reference, we also present these metrics for thresholds maximizing the Youden’s J index (sum of sensitivity and specificity minus one). These include sensitivity, specificity, diagnostic odds ratios (OR), as well as positive (PPV) and negative predictive value (NPV) at 3% prevalence for ATTR-CM or AS, and 1% prevalence for HCM, corresponding to estimated average prevalence of HCM, ATTR-CM, AS in non-randomly selected individuals with age similar to our population and known or suspected cardiovascular disease.^29–33^ Survival analyses were performed using age- and sex-adjusted Cox regression models for all-cause mortality with label-specific output probabilities as an independent covariate split across quintiles. All statistical tests were 2-sided with a significance level of 0·05 unless specified otherwise. Reporting stands consistent with the STROBE (Strengthening the Reporting of Observational Studies in Epidemiology) guidelines.^34^

## RESULTS

### Study population

The development cohort included a total of 10,702 studies with 290,245 echocardiographic videos among 8,460 unique patients across training/validation (7,621 patients with 9,667 studies, median age of 70 [59-79] years, 3,694 [48·5%] female) and testing set (839 patients with 1,035 studies, median age of 71 [61-80] years, 413 [49·2%] female). Across the training, validation, and testing sets, 46,396 (16·0%) videos corresponded to HCM, 8,842 (3·0%) to ATTR-CM and 41,465 (14·3%) to severe AS (**Table 1**).

The YNHHS POCUS cohort included 39,546 studies from 33,127 unique patients (median age 61 [IQR: 45-75 years], 17,276 (52·2%) female) with 78,054 key echocardiographic views (15,751 PLAX, 52,477 PSAX, 9,826 A4c). Compared with the TTE subset, there were relatively more individuals self-identifying as Hispanic (n=5,073 [15·3%] vs n=539 [6·4%], p<0·0001) or Black (8,040 [24·3%] vs 587 [6·9%], *p*<0·0001). Reflecting the prevalence of known (diagnosed) disease, 279 (0·4%) of these videos were in patients with HCM, 172 in patients with ATTR-CM (0·2%), and 1,130 (1·4%) in patients with severe AS.

The MSHS POCUS cohort included 5,906 studies from 5,624 patients (median age of 57 [39-71 years], 1,953 [34·7%] women) with 13,796 clips corresponding to key echocardiographic views (5,876 PLAX, 5,237 PSAX, 2,683 A4c). Among these, 74 (0.5%) videos were in patients with HCM, 32 (0.2%) with ATTR-CM, and 584 (4.1%) with AS (severity unspecified).

### Development of a view-agnostic, multi-label model for HCM and ATTR-CM

On a *study-level* analysis of the held-out testing TTE set, our multi-label view-agnostic classifier successfully discriminated HCM (AUROC 0·95 [95% CI: 0·94-0·96]) and ATTR-CM (0·98 [95% CI: 0·96-0·99]) (**Fig. S1**). On *video-level* comparisons across three key views (PLAX, PSAX [pap.], A4c), the view-agnostic model outperformed view-specific models (δ[AUROC] of +0·05 [95%CI: 0·03-0·07] for HCM & +0·03 [95%CI: 0·01-0·06] for ATTR-CM) (**Fig. S2**). Optimal video-level thresholds for each label based on 90% sensitivity and Youden’s J are presented in **Table S2**.

### Overview of cardiac-focused POCUS studies

As opposed to the standardized protocols of the TTE cohort where the ratio of PLAX:PSAX:A4c views detected by the view classifier was almost 1:1:1, there was a higher prevalence of PSAX views (52,477 [67·2%]), than PLAX (15,751 [20·2%]) or A4c views (9,826 [12·6%]) in the YNHHS POCUS cohort (**Table 1**). Furthermore, the automated view classifier was significantly less confident in assigning a view label (median probability for most likely view class of 0·66 [IQR: 0·45-0·90] vs 0·93 [IQR: 0·69-1·00], *p* <0·0001).

### AI-guided cardiomyopathy detection from real-world ED POCUS videos

On a *video*-level analysis of the YNHHS ED POCUS cohort, our view-agnostic model outperformed view-specific models for HCM (δ[AUROC] of 0·03 [95% CI: 0·00-0·06]) and ATTR-CM (δ[AUROC] of 0·15 [95% CI: 0·10-0·21]). The distribution of the model’s predictions for each distinct label of interest is shown in **Fig. S3**, and demonstrated specificity for each individual condition (HCM, ATTR-CM, AS) despite their overlapping phenotypes.

The diagnostic performance of the AI classifier varied according to the *input view,* the *confidence* of the view classifier, and the *target population* (i.e., when restricting to patients without known heart failure) (AUROC metrics summarized in **Fig. 2 & S4**). For screening of HCM among patients without heart failure history the classifier reached an AUROC of 0·90 [95%CI: 0·80-0·99] when deployed to high-confidence A4c clips (diagnostic odds ratio [OR] of up to 20·5). For ATTR-CM screening, the classifier achieved an AUROC of 0·92 [95%CI: 0·87-0·96] and diagnostic odds ratios of up to 51·5 (see **Tables 2 & S3** for 90% sensitivity and maximal Youden’s J thresholds, respectively). These findings were replicated in the MSHS cohort, where our model achieved an AUROC of 0·89 [95%CI: 0·83-0·94] for HCM and 0·99 [95%CI: 0·99-1·00] for HCM (A4c) and ATTR-CM (PLAX), respectively (**Tables 2 & S3, Fig. S5**).

**Figure 2.**
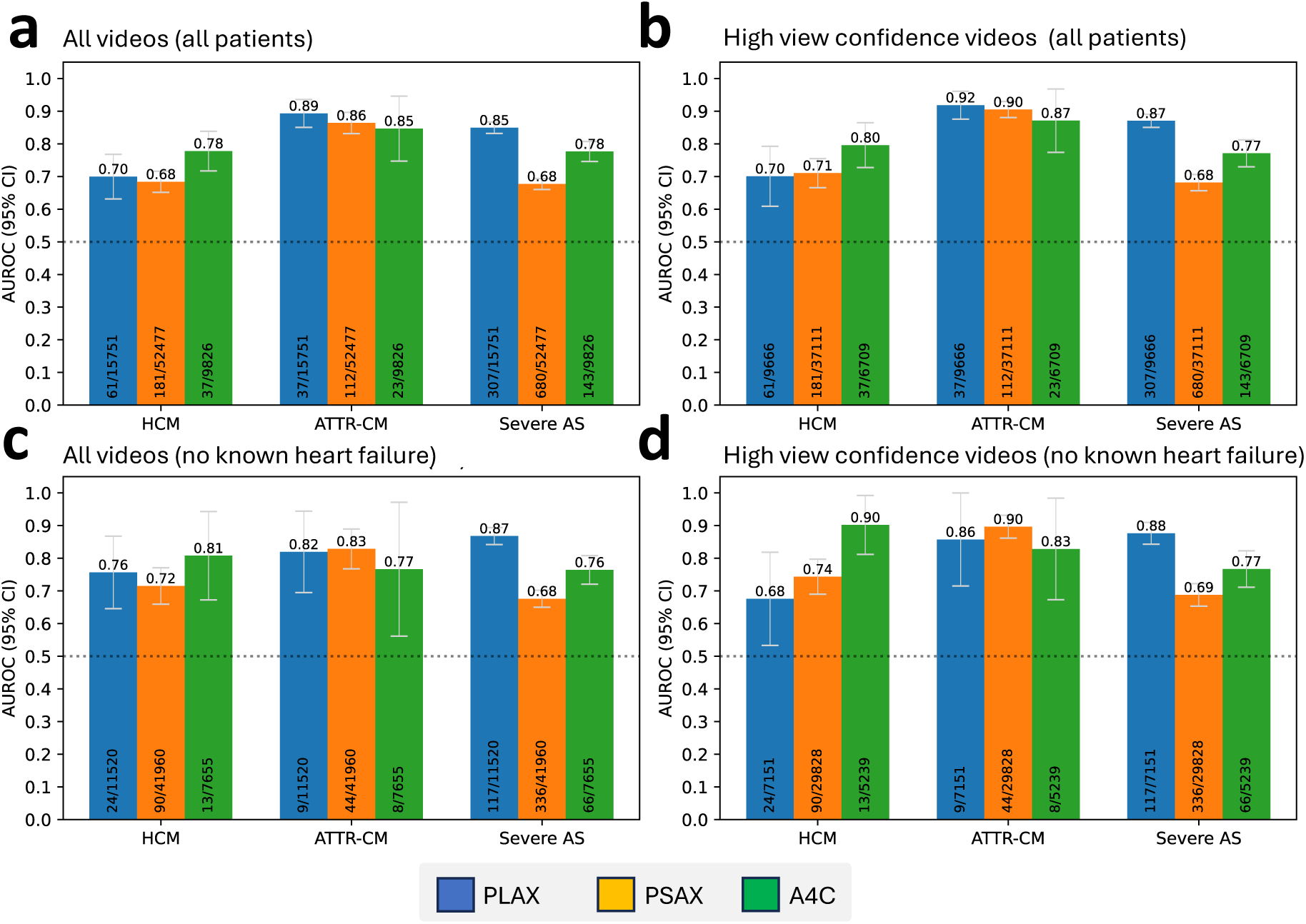
Video-level performance of a view-agnostic multi-label POCUS classifier in YNHHS. Video-level performance (AUROC with 95% CI) for discrimination of HCM, ATTR-CM and severe AS, by deploying a POCUS-adapted model to different echocardiographic views obtained across the emergency rooms of YNHHS (blue = PLAX; orange = PSAX at the papillary muscle level; green = A4C). We present results both for all-comers **(a, b)**, as well as participants without known heart failure at the time of their assessment **(c, d)**, further stratified by the confidence of the automatic view classifier in detecting the corresponding view (all videos **[a, c]** vs view confidence probability of >0.5 **[b, d]**). The numbers at the bottom of each bar denote the counts of cases out of all eligible video counts in this group. All 95% confidence intervals are derived from bootstrapping with 200 replications. AS: aortic stenosis; ATTR-CM: transthyretin amyloid cardiomyopathy; AUROC: area under the receiver operating characteristic curve; CI: confidence interval; HCM: hypertrophic cardiomyopathy; PLAX: parasternal long axis view; POCUS: point-of-care ultrasonography; PSAX: parasternal short axis view; YNHHS: Yale-New Haven Health System.

**Table 2.** Performance of single-view POCUS screening strategies for HCM and ATTR-CM.

| Studies | Label | View | Cohort | AUROC (95%CI) | Sens. | Spec. | PPV* | NPV* | +LR | -LR | Diagn. OR |
| --- | --- | --- | --- | --- | --- | --- | --- | --- | --- | --- | --- |
| All | ATTR-CM | PLAX | YNHHS | 0·89 [0·84, 0·94] | 0·919 | 0·656 | 0·076 | 0·996 | 2·672 | 0·123 | 21·7 |
|  |  |  | MSHS | 0·99 [0·99, 1·00] | 1 | 0·651 | 0·081 | 1 | 2·865 | 0 | n/a |
|  |  | PSAX (PAP) | YNHHS | 0·86 [0·83, 0·90] | 0·857 | 0·747 | 0·095 | 0·994 | 3·387 | 0·191 | 17·7 |
|  |  |  | MSHS | 0·97 [0·96, 0·99] | 1 | 0·613 | 0·074 | 1 | 2·584 | 0 | n/a |
|  | HCM** | A4c | YNHHS | 0·80 [0·63, 0·94] | 0·769 | 0·637 | 0·021 | 0·996 | 2·118 | 0·363 | 5·8 |
|  |  |  | MSHS | 0·89 [0·84, 0·94] | 1 | 0·34 | 0·015 | 1 | 1·515 | 0 | n/a |
| High confidence | ATTR-CM | PLAX | YNHHS | 0·92 [0·87, 0·96] | 0·963 | 0·666 | 0·082 | 0·998 | 2·883 | 0·056 | 51·5 |
|  |  |  | MSHS | 0·99 [0·99, 1·00] | 1 | 0·67 | 0·086 | 1 | 3·03 | 0 | n/a |
|  |  | PSAX (PAP) | YNHHS | 0·91 [0·88, 0·93] | 0·899 | 0·766 | 0·106 | 0·996 | 3·842 | 0·132 | 29·1 |
|  |  |  | MSHS | 0·97 [0·96, 0·98] | 1 | 0·622 | 0·076 | 1 | 2·646 | 0 | n/a |
|  | HCM** | A4c | YNHHS | 0·90 [0·80, 0·99] | 0·889 | 0·656 | 0·025 | 0·998 | 2·584 | 0·169 | 15·3 |
|  |  |  | MSHS | 0·89 [0·83, 0·94] | 1 | 0·368 | 0·016 | 1 | 1·582 | 0 | n/a |

Threshold-dependent metrics are presented at the 90% sensitivity cut-offs. \*PPV and NPV are reported at a simulated prevalence of 3% for ATTR-CM (includes patients with heart failure) and 1% for HCM. \*\*Screening for HCM done in a population without known heart failure in the YNHHS cohort (per prior echocardiography or diagnosis codes). *ATTR-CM: transthyretin amyloid cardiomyopathy; AUROC: area under the receiver operating characteristic curve; HCM: hypertrophic cardiomyopathy; MSHS: Mount Sinai Hospital System; NPV: negative predictive value; OR: odds ratio; PLAX: parasternal long-axis view; PPV: positive predictive value; PSAX (PAP): parasternal short-axis view (papillary muscle); YNHHS: Yale-New Haven Health System.*

### Explainability and representative cases

#### Grad-CAM

Representative Grad-CAM saliency maps for true positive predictions for each label in the YNHHS set are shown in **Fig. 3**. For HCM, these localized to the left ventricle (**Fig. 3a-b**), and for ATTR-CM, the signal localized to the left atrium (**Fig. 3c-d**). For the reference label of severe AS, the focus was on the left ventricle and the aortic valve (when in plane) (**Fig. 3e-f**).

**Figure 3.**
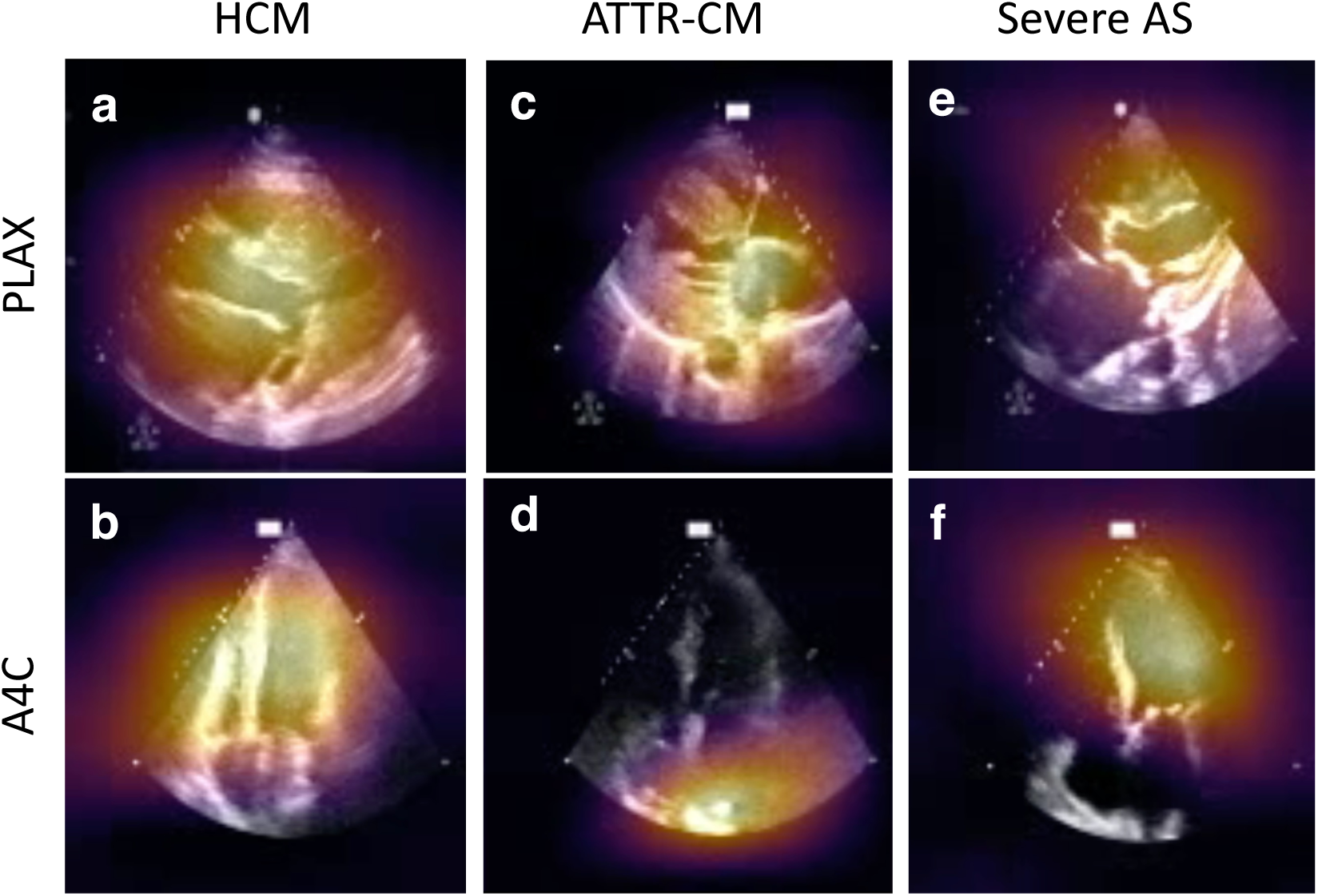
Saliency maps. Activation maps for HCM (**a-b**), ATTR-CM (**c-d**) and AS (**e-f**) across PLAX and A4C views obtained at the point-of-care in the emergency department. A4C: apical-4-chamber view; AS: (severe) aortic stenosis; ATTR-CM: transthyretin amyloid cardiomyopathy; HCM: hypertrophic cardiomyopathy; PLAX: parasternal long axis view.

#### Representative cases

In the **appendix** (**Fig. S6-S7**), we provide an illustrative summary of studies corresponding to the highest and lowest predictions for both cases and controls across all three key views (PLAX, PSAX, A4c). These examples showcase the key challenges of POCUS imaging, including the variation in acquisition quality, probe orientation, off-axis views, and significant noise artifacts.

### Time lag between a positive POCUS screen and clinical diagnosis

To explore the potential of timely screening using AI-assisted POCUS inference, we estimated that among all patients with HCM (n=69) or ATTR-CM (n=48) who had at least one ED POCUS study in the YNHHS (before or after their diagnosis), 40 (58·0%) and 23 (47·9%) had a positive screen by POCUS any time before their eventual confirmatory imaging at the validated 90% sensitivity thresholds of ≥0·07 and 0·10, respectively. Among these participants, the median time between the first positive AI-POCUS and CMR or cardiac scintigraphy was 2·1 [IQR: 0·9-4·5] and 1·9 [IQR: 1·0-3·4] years for HCM and ATTR-CM, respectively (**Fig. 4**).

**Figure 4.**
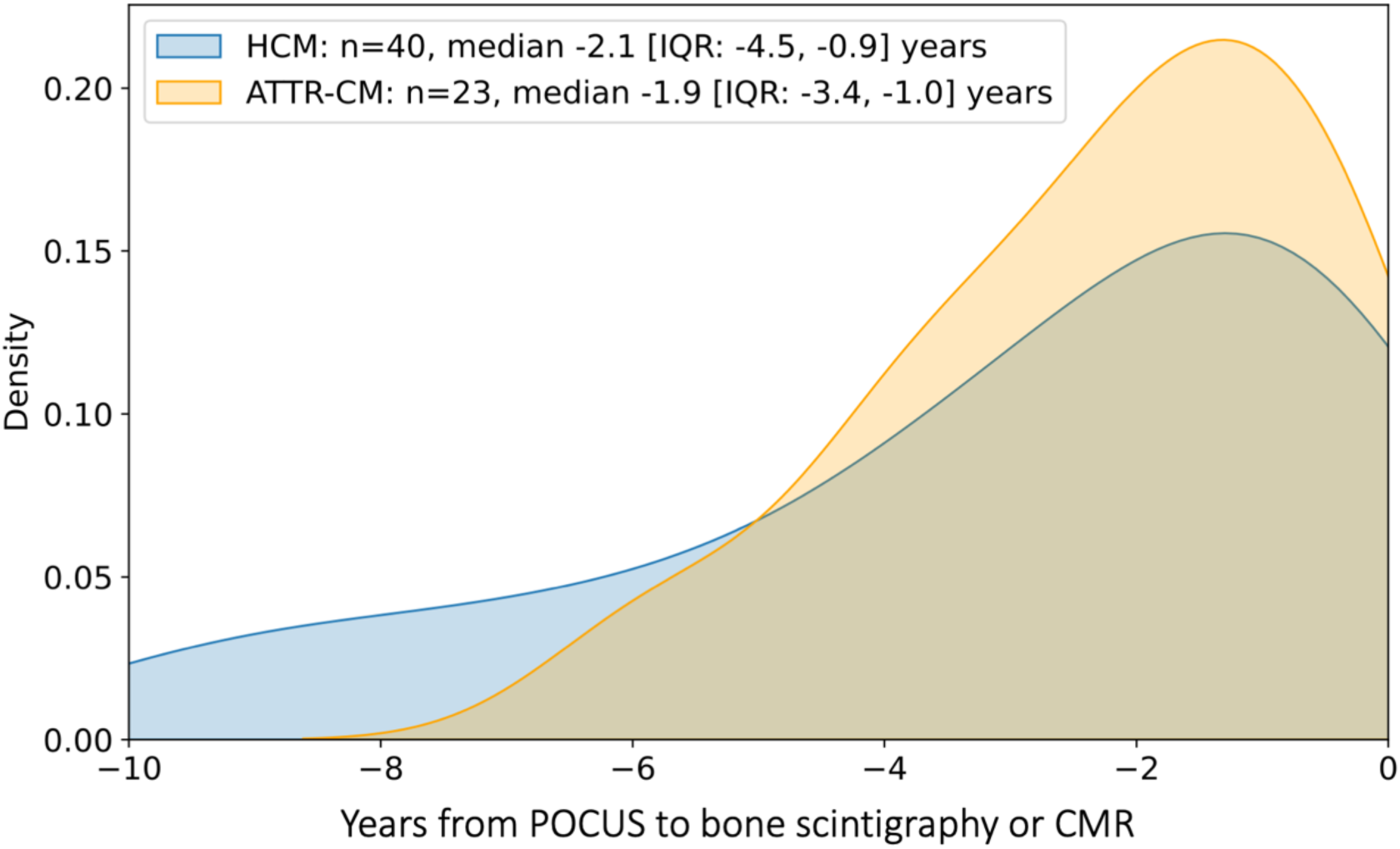
Density plot of time between positive AI-POCUS screen and eventual confirmatory testing. Density plot summarizing the time difference between a positive POCUS screen and confirmatory testing by cardiac magnetic resonance (CMR) or cardiac scintigraphy for 40 and 23 patients with an eventual diagnosis of HCM or ATTR-CM, respectively. ATTR-CM: transthyretin amyloid cardiomyopathy; CMR: cardiac magnetic resonance; HCM: hypertrophic cardiomyopathy; IQR: interquartile range; POCUS: point-of-care ultrasonography.

### AI-POCUS and all-cause mortality among individuals without clinical diagnosis

To address the prognostic implications of potentially under-diagnosed HCM and ATTR-CM, we explored the association between AI-POCUS-defined phenotypes and all-cause mortality among patients who were never diagnosed with cardiomyopathy, including HCM or ATTR-CM. Among 24,448 individuals (median age 58, [IQR 40-73] years; 13,478 [55·1%] women) followed over 2·2 [IQR: 1·1-5·8] years (including a 1-month blanking period), there were 2,738 (11.2%) death events recorded. An HCM or ATTR-CM-like phenotype in the highest (vs lowest) respective quintile conferred a 15% (adj. HR 1·15 [95%CI: 1·02-1·29] for HCM) and 39% higher adjusted risk of mortality (adj. HR 1·39 [95%CI: 1·22, 1·59] for ATTR-CM), respectively (**Fig. 5**).

**Figure 5.**
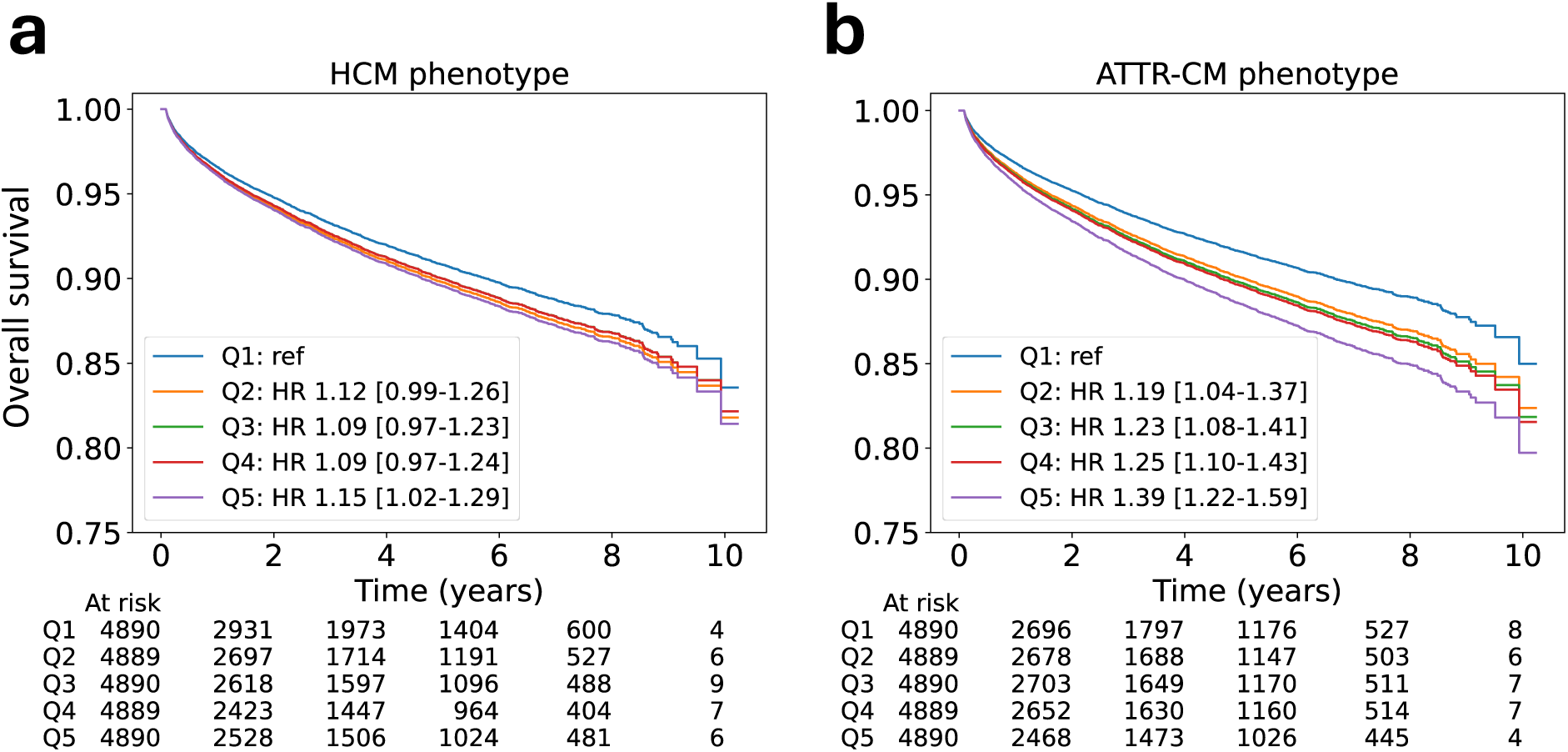
Age- and sex-adjusted survival plots across quintiles of label-specific classifier probabilities. Age- and sex-adjusted survival curves across quintiles (Q1 through Q5) of the output probabilities for HCM **(a)** and ATTR-CM **(b)** on POCUS. Results are presented for individuals who were never diagnosed with cardiomyopathy during the follow-up period (n=24,448). Effect estimates denote HR accompanied by [95% confidence intervals (CI)]. ATTR-CM: transthyretin amyloid cardiomyopathy; CI: confidence interval; HCM: hypertrophic cardiomyopathy; HR: hazard ratio; POCUS: point-of-care ultrasonography.

## DISCUSSION

We demonstrate that an AI algorithm adapted for low-quality cardiac ultrasound acquisitions and off-axis views can reliably identify under-diagnosed cardiomyopathies using simple cardiac POCUS videos. Taken together, our findings suggest a novel role for AI-augmented POCUS interpretation in the opportunistic screening of cardiomyopathies across various settings, expanding the scope of a common and accessible imaging modality that is increasingly available in the community. To achieve this, our approach prompts video-based models to adjust to the unique challenges of handheld cardiac ultrasonography. We demonstrate the generalizability of this approach across video-level analyses of 91,850 distinct POCUS videos from multiple EDs across two large independent healthcare systems for both HCM and ATTR-CM, conditions that are often under-recognized and whose detection requires advanced multimodal imaging. We further provide evidence suggesting that AI-POCUS may enable early diagnosis and that these AI-defined phenotypes have prognostic implications, suggesting the identification of a high-risk group that merits further evaluation and referral for advanced diagnostic testing.

Most currently available AI solutions for echocardiography have been developed for standard transthoracic echocardiography.^14–16,18,19,35–37^ Such models benefit from videos of high quality and anatomical fidelity obtained by certified echocardiography technicians using standardized protocols. Unfortunately, disparities in accessing ambulatory outpatient care do exist and have repeatedly been shown to disproportionately affect marginalized communities.^38–40^ In our study, we found that the proportion of Black and Hispanic individuals was nearly three times higher in POCUS cohort than in the TTE cohort. This highlights the potential to leverage current clinical workflows, particularly for conditions such as HCM and ATTR-CM that benefit from early detection and risk stratification, yet remain under-diagnosed.^8–10^ Moreover, our work suggests a path towards efficient and scalable targeted screening in high-risk communities that may be performed using low-cost equipment, abbreviated protocols, and operators with minimal training.

Our work is both methodologically and clinically innovative. On the methodological front, we describe a scheme that may improve the performance of echocardiography-based AI tools when deployed to real-world POCUS studies by explicitly addressing POCUS-specific challenges as part of the model development process. For instance, we show that view-naïve and synthetic augmentation techniques may help models adjust to various on- and off-axis, high- and low-quality examples. Moreover, a quantifiable surrogate of view confidence allows the algorithm to emphasize examples that deviate from standard views.

On the clinical front, our work directly impacts the scalability of AI-echocardiography tools,^41,42^ ensuring equitable access at the first point of care and offering a platform to reduce disparities arising from differential access to testing. Our proof-of-concept application in a cohort study across temporally and geographically distinct ED visits supports the feasibility of simplified POCUS-based protocols for targeted screening of at-risk individuals in the community. This is further supported by evidence that positive signatures may exist several years before clinical diagnosis of ATTR-CM or HCM and that even among those without the diagnosis, high disease probability is associated with worse long-term outcomes. Together, these highlight the potential of reducing the time from first contact to clinical diagnosis, while also reducing the rates of mis- or under-diagnosis of treatable cardiomyopathies.

Certain limitations merit consideration. First, our study was a retrospective analysis of two large hospital networks, and POCUS studies were performed for individuals presenting to the emergency department with acute symptoms. However, these would represent an appropriate setting for high-risk opportunistic screening. Second, we noted that the performance of the POCUS models was lower than that of TTE. This is likely driven by the inherently lower signal-to-noise ratio of POCUS acquisitions, which can potentially be improved by emerging technologies that enable assisted acquisition. Our evaluation of the model performance is further challenged by the known under-diagnosis of HCM and ATTR-CM, with several cases likely flagged as false positives in our study. This is demonstrated by many individuals flagging years before their clinical diagnosis and with an elevated risk of death. Both of these may introduce a degree of ascertainment bias that might suggest lower than actual performance of the model.

Finally, diagnoses in the MSHS cohort were adjudicated based on ICD codes, which can have limited accuracy and sensitivity in flagging key conditions. Moreover, the case volumes were lower. However, our models retained consistent performance across institutions despite these differences, further supporting the specificity of our approach that appears to be independent of the exact approach to defining the labels.

## CONCLUSIONS

We demonstrate that AI models adapted for use with POCUS can reliably identify key under-diagnosed, yet increasingly treatable, cardiomyopathies directly on simple, low-quality cardiac ultrasound. We show the potential of a novel, scalable, and inexpensive screening approach that uses automated AI-based inference on an accessible and portable modality to identify conditions that typically require advanced multimodality imaging, with the potential for early diagnosis and improved patient outcomes.

## Data Availability

All data produced in the present study are available upon reasonable request to the authors.

## Data Sharing Agreement

The underlying videos may contain identifiable information and cannot be released at this time. The analytical code can be made available upon reasonable request to the authors.

