## Appendix for "Artificial intelligence-guided detection of under-recognized cardiomyopathies on point-of-care cardiac ultrasound: a multi-center study"

**Appendix (Online Supplement)**

Evangelos K. Oikonomou MD[a], Akhil Vaid MD[b,c], Gregory Holste BA[d], Andreas Coppi PhD[e], Robert L. McNamara MD[a], Cristiana Baloescu MD[f], Harlan M. Krumholz MD[a,e], Zhangyang Wang PhD[d], Donald J. Apakama MD[g], Girish N. Nadkarni MD[b,c], Rohan Khera MD[a,e,h,i,j]*

[a] Section of Cardiovascular Medicine, Department of Internal Medicine, Yale School of Medicine, New Haven, CT, USA
[b] The Charles Bronfman Institute for Personalized Medicine, Icahn School of Medicine at Mount Sinai, New York, NY, USA.
[c] The Division of Data Driven and Digital Medicine, Department of Medicine, Icahn School of Medicine at Mount Sinai, New York, NY, USA.
[d] Department of Electrical and Computer Engineering, The University of Texas at Austin, Austin, TX, USA
[e] Center for Outcomes Research and Evaluation, Yale-New Haven Hospital, New Haven, CT, USA
[f] Department of Emergency Medicine, Yale School of Medicine, New Haven, CT, USA
[g] Department of Emergency Medicine, Icahn School of Medicine at Mount Sinai, New York, New York, USA
[h] Department of Biostatistics, Yale School of Public Health, New Haven, CT, USA
[i] Section of Biomedical Informatics and Data Science, Yale School of Medicine, New Haven, CT, USA
[j] Section of Health Informatics, Department of Biostatistics, Yale School of Public Health, New Haven, CT, USA

**Table of Contents**

**Page 2-4:** Supplemental Methods
**Pages 5-7:** Supplemental Tables S1-S3
**Pages 8-14:** Supplemental Figures S1-S7
**Page 15:** Supplemental References

***Corresponding author:**
Rohan Khera, MD, MS
195 Church St, 6th Floor, New Haven, CT 06510
203-764-5885;; @rohan_khera

### Supplemental Methods

#### Definitions of cases and controls in the Yale-New Haven Health System (YNHHS)

***HCM***: All individuals with an ICD-9/ICD-10 code for any cardiomyopathy (425, I42.0, I42.1, I42.2, I42.5, I42.8, I42.9, I43.1, I43.8) or heart failure (428, I50*), inclusive of HCM-specific ICD-9/10 codes (425.1, 425.11, 425.18, I42.1, I42.2) were identified. To maximize the specificity of our definition, given the known unreliability of billing/administrative codes in accurately capturing patient phenotypes,[1] we required those with positive labels for this condition to have undergone cardiac magnetic resonance (CMR) imaging with the final interpretation/conclusion supporting the presence of the diagnosis.[2] Given that HCM is a genetic cardiomyopathy, we included all available echocardiograms regardless of their timing relative to the time of diagnosis.[3]

***Amyloid cardiomyopathy (ATTR-CM)***: We screened for all individuals with an ICD-based diagnosis of any cardiomyopathy or heart failure as above or an amyloidosis-specific code (277.3, 277.30, 277.39, E85.2, E85.82, E85.4, E85.8, E85.9, excluding E85.81 [light chain amyloidosis]). Similar to HCM, to increase the specificity of the label, we required positive labels to have undergone bone scintigraphy (with $Tc^{99m}$-pyrophosphate [PYP]), which was interpreted as positive for cardiac uptake by the interpreting physician (i.e., a semi-quantitative visual score of 2 or 3 or heart to contralateral lung ratio >1.5).[4] For positive cases, we defined the time of diagnosis as the time of the positive PYP scan, and, to account for the delay between disease onset and diagnosis (median delay of ~13 months as previously reported in the literature),[5] we included echocardiograms performed up to 12 months before this date (and any time after).

***Controls:*** These were defined by randomly sampling echocardiograms from the same period, after excluding any positive HCM or ATTR-CM cases, and after excluding intermediate phenotypes (i.e., CMR findings suggestive of possible HCM, or equivocal PYP results). The study sample was further enriched for cases of severe AS, including severe low-flow, low-gradient AS, based on the interpretation of a TTE exam by a board-certified reader and in agreement with existing guidelines.[6,7] This was done to ensure the model learned to identify AS, a separate pathology, rather than a confounder of ATTR-CM or other cardiomyopathies.

#### Automated view characterization and alignment assessment

We implemented our previously published end-to-end pre-processing pipeline for echocardiographic studies stored in DICOM format, which involves loading the pixel data, masking out pixels in the periphery to remove identifying information and converting to Audio Video Interleave (.AVI) format.[19] We then randomly sampled ten frames from each video, down-sampled to 224x224 pixels, and fed these frames through a previously validated VGG19 convolutional neural network (CNN) that enables video-level classification of 18 echocardiographic views by assigning a probability that a given video corresponds to a standard anatomical view (with probabilities adding up to 1 across all views).[26] A predicted view was then assigned based on the view class that has the highest probability. The highest probability value (0-1) is then used to define a metric of anatomical alignment with standard echocardiographic views. In other words, greater anatomical correctness and view quality were associated with higher confidence in the CNN model's output. Next, we performed more thorough cleaning and de-identification by binarizing each video frame with a fixed threshold, masking out all pixels outside the convex hull of the largest contour, and down-sampling to 112x112 pixels, as described in our previous work.[19,27]

**Model customization (extended)**
***Natural and synthetic data augmentation methods*:** We trained both separate models for each key views-of-interest, namely PLAX, PSAX, and A4C, followed by all-inclusive, view-naïve models trained in pooled datasets that included all parasternal (long and short) and apical views with the classifier blinded to the input view. This enabled a head-to-head comparison of how view-specific versus view-agnostic approaches generalize to real-world POCUS acquisitions. We further applied a series of data augmentations to account for variable orientation and off-axis views that included random zero padding by up to 8 pixels in each spatial dimension, random horizontal flipping with (probability 0.5), and a random rotation within -10 and 10 degrees (probability 0.5). After augmentation, each video clip's intensities were normalized to 0-1 and standardized using the channel-wise means and standard deviations from the Kinetics-400 training dataset.

***Quality-adjusted weights and loss function:*** We defined a loss function that encouraged the model to learn from lower-quality cases. We took the sigmoid binary cross entropy (BCE) loss function (implemented with PyTorch's *BCEWithLogitsLoss*), and incorporated both label-specific weights to account for rare labels,[29] as well as inverse weighting based on the view alignment probabilities (see ***Supplement***).[26] Higher probabilities (i.e., PLAX view probability of 1.00) denote greater anatomical correctness compared with lower probabilities. The aim of this weighting scheme was to penalize the model for missing under-represented labels, especially in the context of a challenging view. We also applied label smoothing ($\alpha$=0.1) to help regularize the model and penalize overconfidence in its predictions.[30,31]

**Customized loss function description:**

**Step 1: View Weights:**
For each sample *i* in the batch:

$$V_i = \frac{1}{(P_\mathrm{i} + 10^{-5})^2}$$

*Where $P_i$ is the view probability for sample i.*

These weights are then normalized across the batch:

$$V_i' = \frac{Vi}{\sum_{j=1}^{N} V_j}$$

*Where N is the number of samples in the batch.*

**Step 2: Class Weights:**
For each label *k* in a sample *i:*

$$C_{ik} = T_{ik} \times W_{1k} + (1 - T_{ik}) \times W_{0k}$$

*Where $T_{ik}$ is the true binary label for label k in sample i, and $W_{1k}$ and $W_{0k}$ are the class weights for the positive and negative classes of label k, respectively.*

**Step 3: Combined Weights:**
The combined weight for each label in each sample is the product of the view weight and the class weight, normalized across the batch:

$$W_{ik} = \frac{C_{ik} \; x \; V_i'}{\sum_{j=1}^{N} \sum_{k=1}^{K} C_{jk} \times V_j'}$$

*Where K is the number of labels.*

**Step 4: Weighted Loss:**
Finally, the weighted loss for the batch is the sum of the individual weighted losses:

$$Weighted\ loss = \sum_{i=1}^{N} \sum_{i=1}^{K} L_{ik} \times W_{ik}$$

*Where $L_{ik}$ is the binary cross-entropy loss for label k in sample i.*

**Model training (extended)**
Models were trained on four NVIDIA Tesla T4 GPUs with the Adam optimizer, a learning rate of $10^{-4}$, a batch size of 56 to maximize GPU utilization, and a random dropout of 0.25. Each model was trained with randomly sampled video clips of 16 frames and sampling one out of every five frames to enable a global capture of the cardiac cycle (median number of frames 61 $25^{th}$-$75^{th}$ percentile: 50-85]). We applied optional padding with empty frames along the temporal axis if either the video was too short or the randomly chosen starting point of the clip was near the end of the video.

**Key packages used**
Key analyses were performed using Python 3.9.7, using pytorch 1.8.0, torchvision 0.9.0, and scipy 1.7.3.

### Supplemental Tables

#### Table S1. ICD diagnosis and procedures codes for inclusion and exclusion criteria.

| | |
|---|---|
| **Conditions to exclude from POCUS analysis** | |
| **End-stage renal disease** | "N18.6", "Z99.2", "I12.0", "585.6", "V45.11" |
| **Heart transplant** | "I25.7", "I25.811", "I25.812", "T86.20", "T86.21", "T86.22", "T86.23", "T86.298", "T86.31", "T86.39, "Z48.21", "Z76.82", "Z94.1", "Z94.3" |
| **Aortic valve replacement** | "02RF", "35.21", "35.22" |
| **Condition-defining labels in the MSHS** | |
| **HCM** | "I42.1" (obstructive), "I42.2",<br>"425.1", "425.11" (obstructive), "425.18" |
| **Cardiomyopathy** | "I420", "I421", "I422", "I425", "I428", "I429", "I431", "I438", "425" |
| **Heart failure** | “I11.0”, “I13.0”, “I13.2”, “I50”, “I50.0”, “I50.1”, “I50.9”, “428”, “428.0”, “428.1”, “428.9” |
| **Transthyretin amyloidosis + cardiomyopathy** | "E85.2", "E85.82" (in combination with cardiomyopathy or heart failure code) |
| **Aortic stenosis (defined by diagnosis codes, severity unspecified)** | "I35.0", "I35.2", "I06.0", "I06.2" |

ATTR: transthyretin amyloidosis; HCM: hypertrophic cardiomyopathy; ICD: International Classification of Diseases; MSHS: Mt Sinai health system; POCUS: point-of-care ultrasound. ICD codes contain ICD-9, ICD-10 codes.

**Table S2. Video-level performance metrics across key thresholds in the TTE testing set.**

| Label | Criterion | Threshold | Sensitivity | Specificity |
|---|---|---|---|---|
| HCM | Youden's J | 0.372 | 0.743 | 0.825 |
| | 90% Sensitivity | 0.070 | - | 0.556 |
| ATTR-CM | Youden's J | 0.262 | 0.881 | 0.899 |
| | 90% Sensitivity | 0.100 | - | 0.839 |
| Severe AS | Youden's J | 0.519 | 0.759 | 0.780 |
| | 90% Sensitivity | 0.249 | - | 0.571 |

AS: aortic stenosis; ATTR-CM: transthyretin amyloid cardiomyopathy; HCM: hypertrophic cardiomyopathy.

**Table S3 | Performance of single-view POCUS screening strategies for HCM and ATTR-CM.**

| Studies | Label | View | Cohort | AUROC (95%CI) | Sens. | Spec. | PPV* | NPV* | +LR | -LR | Diagn. OR |
|---|---|---|---|---|---|---|---|---|---|---|---|
| All | ATTR-CM | PLAX | YNHHS | **0·89 [0·84, 0·94]** | 0·811 | 0·765 | 0·096 | 0·992 | 3·451 | 0·247 | 14·0 |
| | | | MSHS | **0·99 [0·99, 1·00]** | 1 | 0·794 | 0·131 | 1 | 4·854 | 0 | n/a |
| | | PSAX (PAP) | YNHHS | **0·86 [0·83, 0·90]** | 0·679 | 0·862 | 0·132 | 0·989 | 4·92 | 0·372 | 13·2 |
| | | | MSHS | **0·97 [0·96, 0·99]** | 1 | 0·765 | 0·116 | 1 | 4·255 | 0 | n/a |
| | HCM** | A4c | YNHHS | **0·80 [0·63, 0·94]** | 0·538 | 0·902 | 0·053 | 0·995 | 5·49 | 0·512 | 10·7 |
| | | | MSHS | **0·89 [0·84, 0·94]** | 0·909 | 0·726 | 0·032 | 0·999 | 3·318 | 0·125 | 26·5 |
| High confidence | ATTR-CM | PLAX | YNHHS | **0·92 [0·87, 0·96]** | 0·852 | 0·77 | 0·103 | 0·994 | 3·704 | 0·192 | 19·3 |
| | | | MSHS | **0·99 [0·99, 1·00]** | 1 | 0·802 | 0·135 | 1 | 5·051 | 0 | n/a |
| | | PSAX (PAP) | YNHHS | **0·91 [0·88, 0·93]** | 0·734 | 0·874 | 0·153 | 0·991 | 5·825 | 0·304 | 19·2 |
| | | | MSHS | **0·97 [0·96, 0·98]** | 1 | 0·765 | 0·116 | 1 | 4·255 | 0 | n/a |
| | HCM** | A4c | YNHHS | **0·90 [0·80, 0·99]** | 0·667 | 0·911 | 0·07 | 0·996 | 7·494 | 0·366 | 20·5 |
| | | | MSHS | **0·89 [0·83, 0·94]** | 0·900 | 0·734 | 0·033 | 0·999 | 3·383 | 0·136 | 24·9 |

Threshold-dependent metrics are presented at cut-off that maximizes Youden's J. *PPV and NPV are reported at a simulated prevalence of 3% for ATTR-CM (includes patients with heart failure) and 1% for HCM. **Screening for HCM done in a population without known heart failure in the YNHHS cohort (per prior echocardiography or diagnosis codes). *ATTR-CM: transthyretin amyloid cardiomyopathy; AUROC: area under the receiver operating characteristic curve; HCM: hypertrophic cardiomyopathy; MSHS: Mount Sinai Hospital System; NPV: negative predictive value; OR: odds ratio; PLAX: parasternal long-axis view; PPV: positive predictive value; PSAX (PAP): parasternal short-axis view (papillary muscle); YNHHS: Yale-New Haven Health System.*

### Supplemental Figures

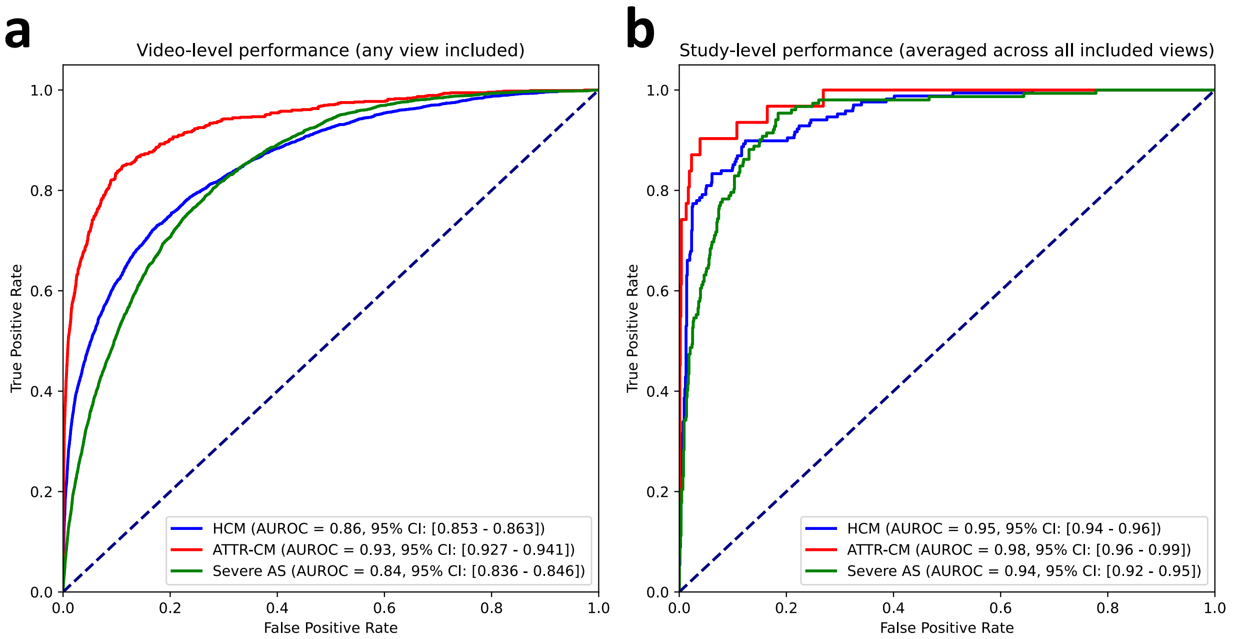


**Figure S1 | Video and study-level performance of a view-agnostic multi-label deep learning classifier. (a)** Video-level performance (across all available parasternal long, parasternal short and apical views) for discrimination of HCM, ATTR-CM and AS. **(b)** Study-level performance by arithmetic mean averaging of all available videos within a given study. All 95% confidence intervals are derived from bootstrapping with 200 replications.AS: (severe) aortic stenosis; ATTR-CM: transthyretin amyloid cardiomyopathy; AUROC: area under the receiver operating characteristic curve; CI: confidence interval; HCM: hypertrophic cardiomyopathy.

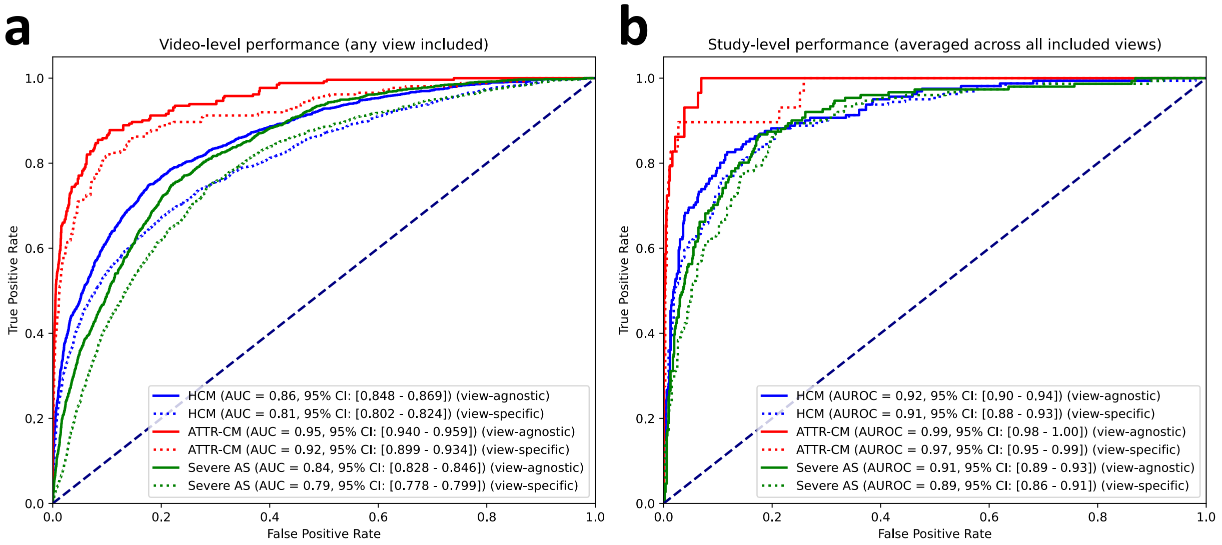


**Figure S2 | Head-to-head video-level and study-level performance of a view-agnostic vs view-specific multi-label deep learning algorithm. (a)** Video-level performance (across all available PLAX, PSAX [papillary muscle level] and A4C views) for discrimination of HCM (blue), ATTR-CM (red) and AS (green) using view-agnostic (uninterrupted line) vs view-specific models (dotted lines). The graphs compare the performance of a master algorithm (view-specific) trained across all views against the selective deployment of algorithms trained with videos from specific views (view-agnostic) **(b)** Study-level performance by simple mean averaging of all available videos within a given study. All 95% confidence intervals are derived from bootstrapping with 200 replications. A4C: apical-4-chamber view; AS: (severe) aortic stenosis; ATTR-CM: amyloid transthyretin cardiomyopathy; AUROC: area under the receiver operating characteristic curve; CI: confidence interval; HCM: hypertrophic cardiomyopathy; PLAX: parasternal long axis view; PSAX: parasternal short axis view.

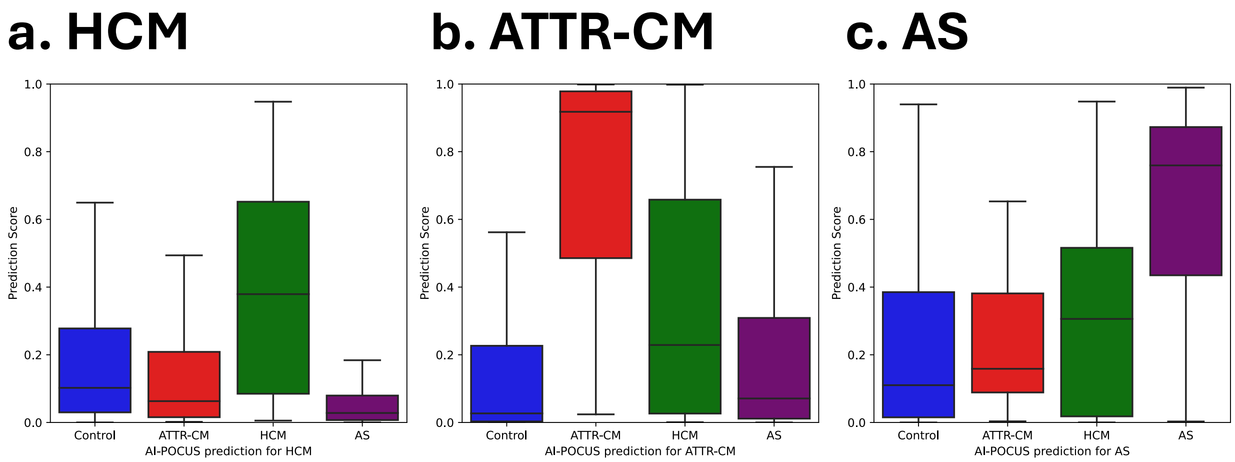


**Figure S3 | Distribution of label-specific predictions for a view agnostic-model.** Box-and-whisker plots denoting the median (horizontal line), interquartile range (box length) and $10^{th}$ to $90^{th}$ percentile (whiskers) for the output of a view-agnostic, multilabel classifier for **(a)** HCM, **(b)** ATTR-CM, **(c)** severe AS, stratified by the underlying label for ATTR-CM, HCM and AS, respectively. AS: aortic stenosis; ATTR-CM: amyloid transthyretin cardiomyopathy; HCM: hypertrophic cardiomyopathy; POCUS: point-of-care ultrasonography.

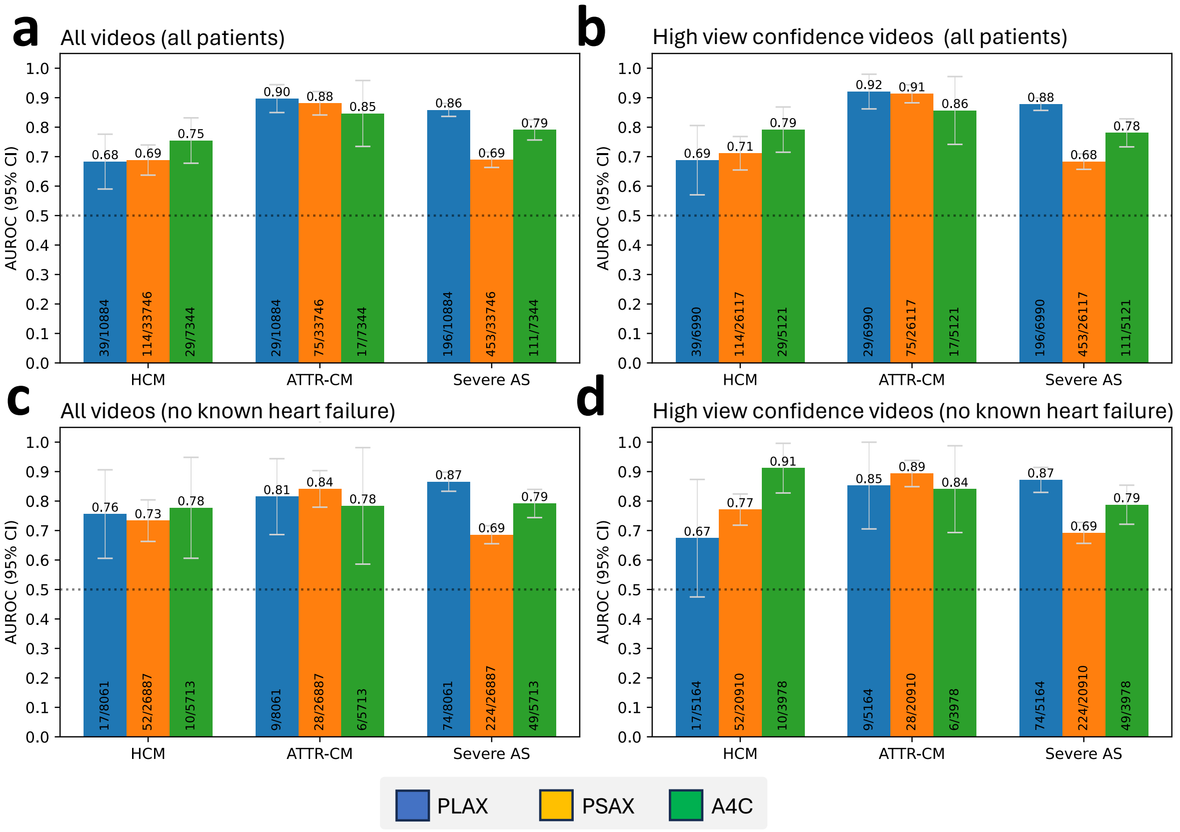


**Figure S4 | Study-level performance of a view-agnostic multi-label POCUS classifier in YNHHS.** Study-level performance (AUROC with 95% CI) for discrimination of HCM, ATTR-CM and severe AS, by deploying a POCUS-adapted, view-agnostic model to different echocardiographic views obtained across the emergency rooms of YNHHS (blue = PLAX; orange = PSAX at the papillary muscle level; green = A4C). We present results both for all-comers **(a, b)**, as well as participants without known heart failure at the time of their assessment **(c, d)**, further stratified by the confidence of the automatic view classifier in picking up the anatomical correctness of the view (all videos **[a, c]** vs view confidence probability of >0.5 **[b, d]**). The numbers at the bottom of each bar denote the counts of cases out of all eligible study counts in this group. All 95% confidence intervals are derived from bootstrapping with 200 replications. AUROC: area under the receiver operating characteristic curve; CI: confidence interval; HCM: hypertrophic cardiomyopathy; PLAX: parasternal long axis view; POCUS: point-of-care ultrasonography; PSAX: parasternal short axis view; YNHHS: Yale-New Haven Health System.

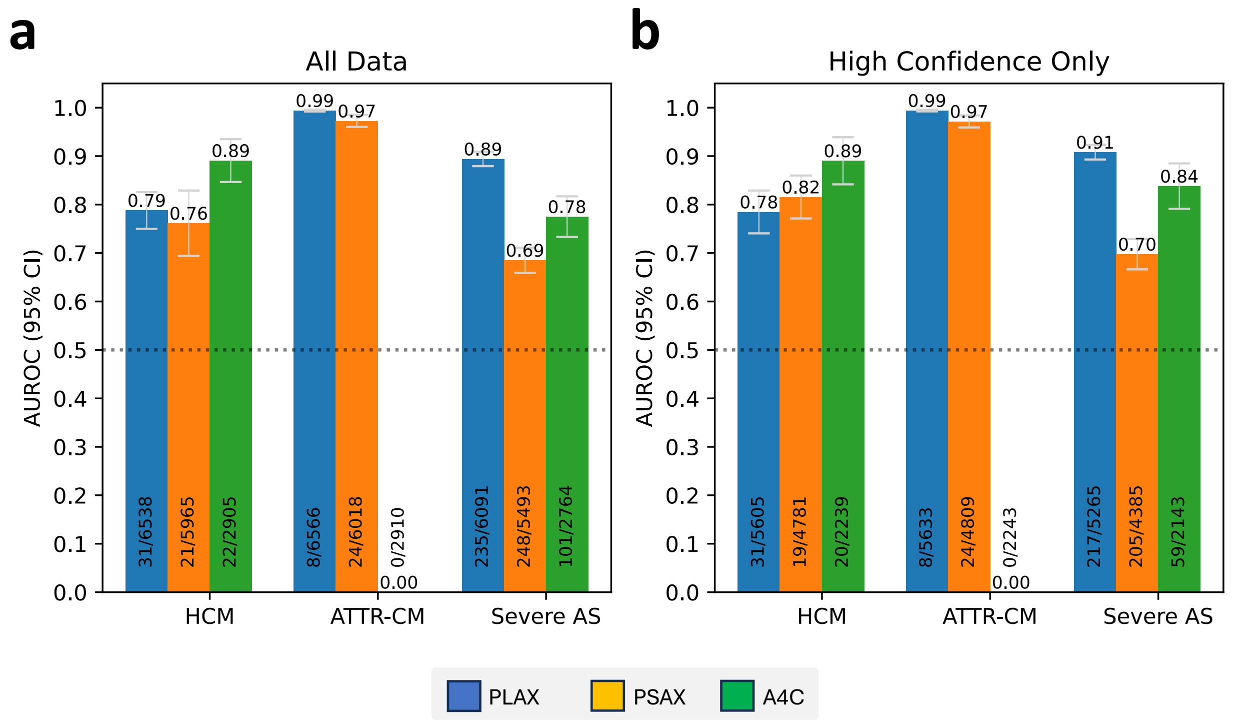


**Figure S5 | Video-level performance of a view-agnostic multi-label POCUS classifier in MSHS.** Video-level performance (AUROC with 95% CI) for discrimination of HCM, ATTR-CM and severe AS, by deploying a POCUS-adapted, view-agnostic model to different echocardiographic views obtained in the emergency department in the MSHS system (blue = PLAX; orange = PSAX at the papillary muscle level; green = A4C). We present results both all videos **(a)** as well as videos where the view was classified with high confidence **(b)**. The numbers at the bottom of each bar denote the counts of cases out of all eligible video counts in this group. All 95% confidence intervals are derived from bootstrapping with 200 replications. AUROC: area under the receiver operating characteristic curve; CI: confidence interval; HCM: hypertrophic cardiomyopathy; LVH: left ventricular hypertrophy; MSHS: Mt Sinai Health System; PLAX: parasternal long axis view; POCUS: point-of-care ultrasonography; PSAX: parasternal short axis view.

**Figures S6-S7 | Sample frames from the highest and lowest predictions for each label across key views in the YNHHS POCUS cohort.** Representative frames of the highest and lowest predictions for each label across key views in the POCUS cohort. A4C: apical-4-chamber view; ATTR-CM: transthyretin amyloid cardiomyopathy; HCM: hypertrophic cardiomyopathy; PLAX: parasternal long axis view; POCUS: point-of-care ultrasound; PSAX: parasternal short axis view.

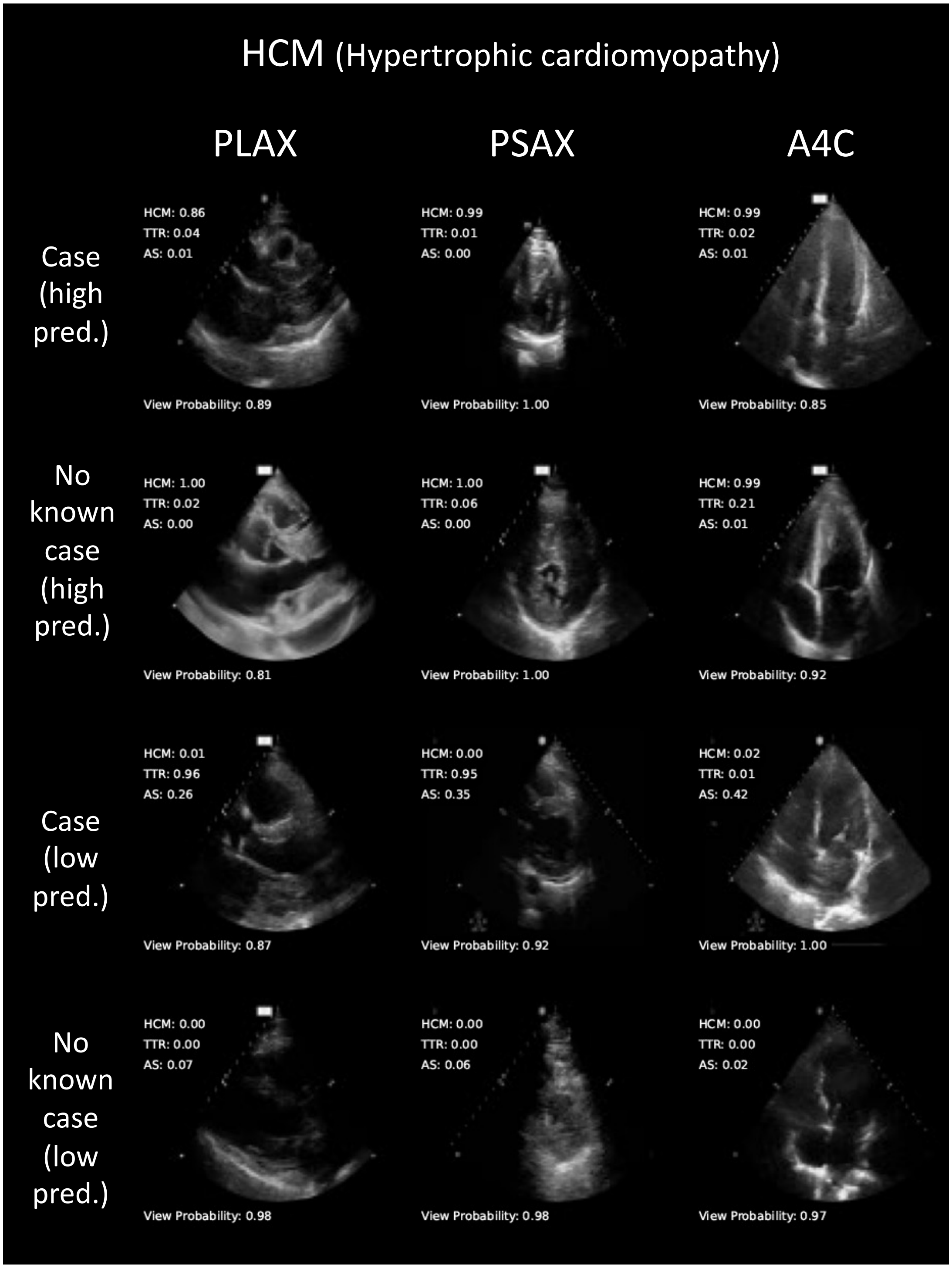

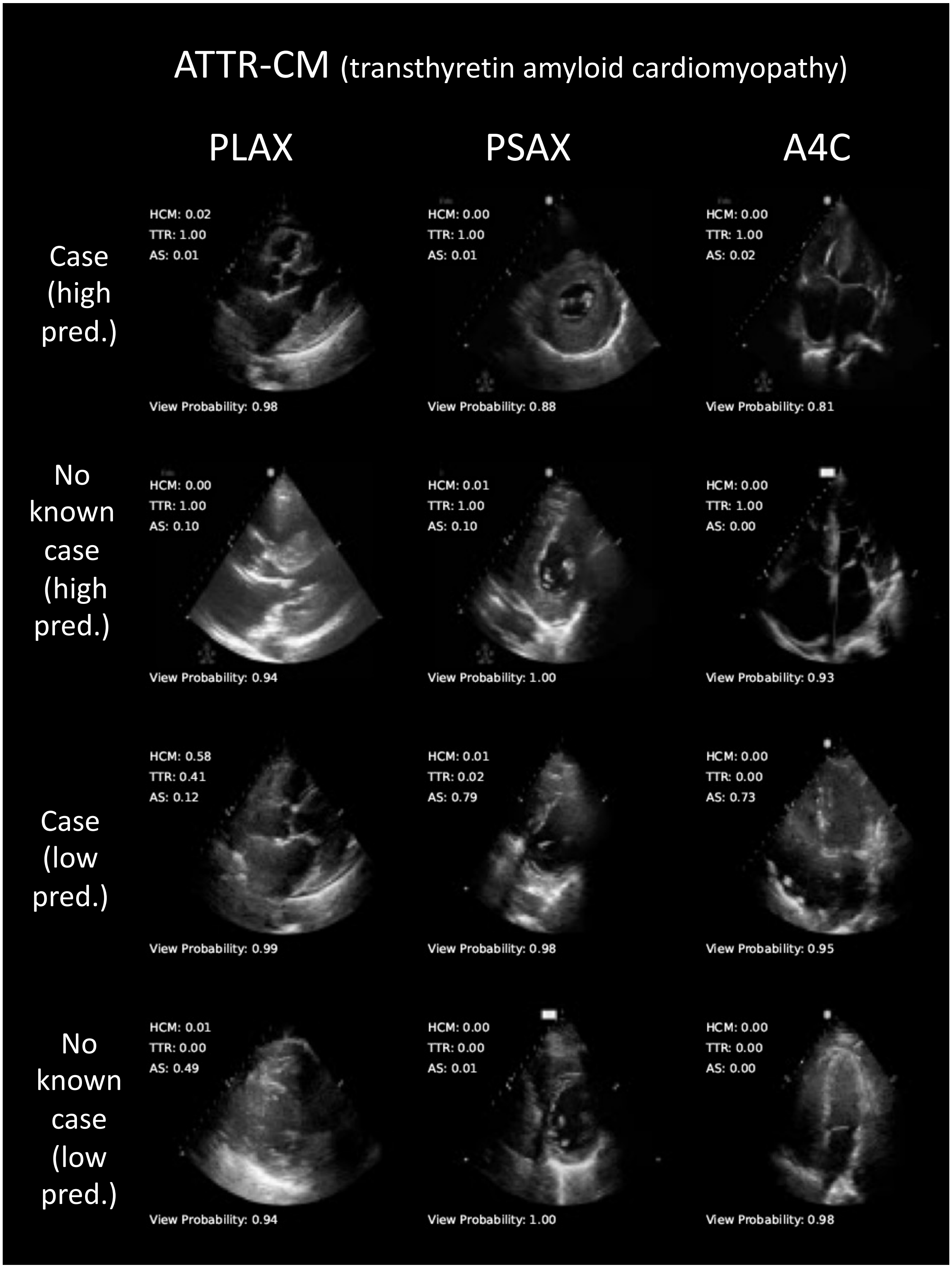
ATTR-CM (transthyretin amyloid cardiomyopathy)
PLAX
PSAX
A4C
Case (high pred.)
HCM: 0.02
TTR: 1.00
AS: 0.01
View Probability: 0.98
HCM: 0.00
TTR: 1.00
AS: 0.01
View Probability: 0.88
HCM: 0.00
TTR: 1.00
AS: 0.02
View Probability: 0.81
No known case (high pred.)
HCM: 0.00
TTR: 1.00
AS: 0.10
View Probability: 0.94
HCM: 0.01
TTR: 1.00
AS: 0.10
View Probability: 1.00
HCM: 0.00
TTR: 1.00
AS: 0.00
View Probability: 0.93
Case (low pred.)
HCM: 0.58
TTR: 0.41
AS: 0.12
View Probability: 0.99
HCM: 0.01
TTR: 0.02
AS: 0.79
View Probability: 0.98
HCM: 0.00
TTR: 0.00
AS: 0.73
View Probability: 0.95
No known case (low pred.)
HCM: 0.01
TTR: 0.00
AS: 0.49
View Probability: 0.94
HCM: 0.00
TTR: 0.00
AS: 0.01
View Probability: 1.00
HCM: 0.00
TTR: 0.00
AS: 0.00
View Probability: 0.98
